# Appetitive mapping of the human nucleus accumbens

**DOI:** 10.1101/2022.09.12.22279834

**Authors:** Jonathon J. Parker, Cammie E. Rolle, Rajat S. Shivacharan, Daniel A. N. Barbosa, Austin Feng, Yuhao Huang, Bina W. Kakusa, Tom Prieto, Richard A. Jaffe, Nolan R. Williams, Casey H. Halpern

## Abstract

There is strong evidence for the putative role of the nucleus accumbens (NAc) in appetitive motivation. A first-in-human feasibility trial of responsive deep brain stimulation (rDBS) for loss of control eating provided a rare opportunity to examine the physiologic and structural underpinnings of NAc function in a human participant with dysregulated appetition. Patient-specific probabilistic tractography was supplemented by intraoperative microelectrode recordings and stimulation testing to confirm appetitive circuit engagement. Personalized visual stimuli we used to provoke and map appetitive units within the NAc, prior to the surgical implantation of a rDBS system, which was safe, feasible, and well-tolerated. Ambulatory patient-triggered recordings provide ongoing electrophysiologic surveillance of NAc activity time-locked to eating behavior in the real-world. This technique described provides a proof-of-concept for utilizing simultaneous intracranial activity and real-time appetitive responses to guide implantation of a rDBS system to treat loss of control eating.

---

Bariatric surgery is the most effective treatment for refractory obesity but can be complicated by comorbid binge eating disorder (BED), which manifests as frequent bouts of compulsive, loss of control (LOC) eating (1–4). Given the episodic nature of LOC eating, an on-demand, neuromodulatory approach was conceived previously in mice, automated by anticipatory local field potential (LFP) recordings to facilitate a closed-loop or responsive DBS (rDBS) strategy (5). This body of work provided the basis to initiate a first-in-human clinical trial of NAc rDBS in patients with treatment-refractory, morbid obesity exhibiting LOC eating (ClinicalTrials.gov NCT03868670) (6,7). Structural neuroimaging, subject-specific tractography, and a novel awake electrophysiologic protocol were used to map the human NAc region prior to and during the implantation of the first participant with an rDBS system. Building on our prior work mapping a contamination obsession in a patient with obsessive-compulsive disorder, this conceptual framework was applied to design a personalized method to map appetitive units (8).

We report the implementation of this mapping protocol including 1) personalized tractography, 2) intraoperative patient-specific awake appetitive testing, and 3) stimulation testing to evoke a positive affect. The final mapping criterion arose from our hypothesis that a timely, evoked positive affect could be therapeutic for LOC eating, which is often preceded by pre-meal negative affect (9,10). Notably, given this first-ever surgery was part of an early feasibility assessment of bilateral NAc rDBS, *a priori* criteria were established to require that two of the three mapping strategies were successful unilaterally.

Targets within the NAc region were chosen using published coordinates for guidance of lead trajectories (5,11). As recently highlighted (12–14), patient-specific tractography may have utility in predicting response to DBS. Thus, we defined a patient-specific subdivision of the NAc region, which was hypothesized to contain projections to the lateral hypothalamus (LH), a circuit involved in hedonic eating behavior, to refine targeting (Figure 1A) (11,15–18). Appetitive stimuli (n=26 total images) with patient-specific appetitive ratings greater than 8 (using a 10-point Likert scale; mean +/- SD, 7.9 +/- 2.4) were selected as our clinical instrument for intraoperative provocation. The provocation paradigm included instruction, fixation/anticipation, and presentation phases with pixel-wise photo scrambles generated for color and brightness stimuli control (19).

**Figure 1.**
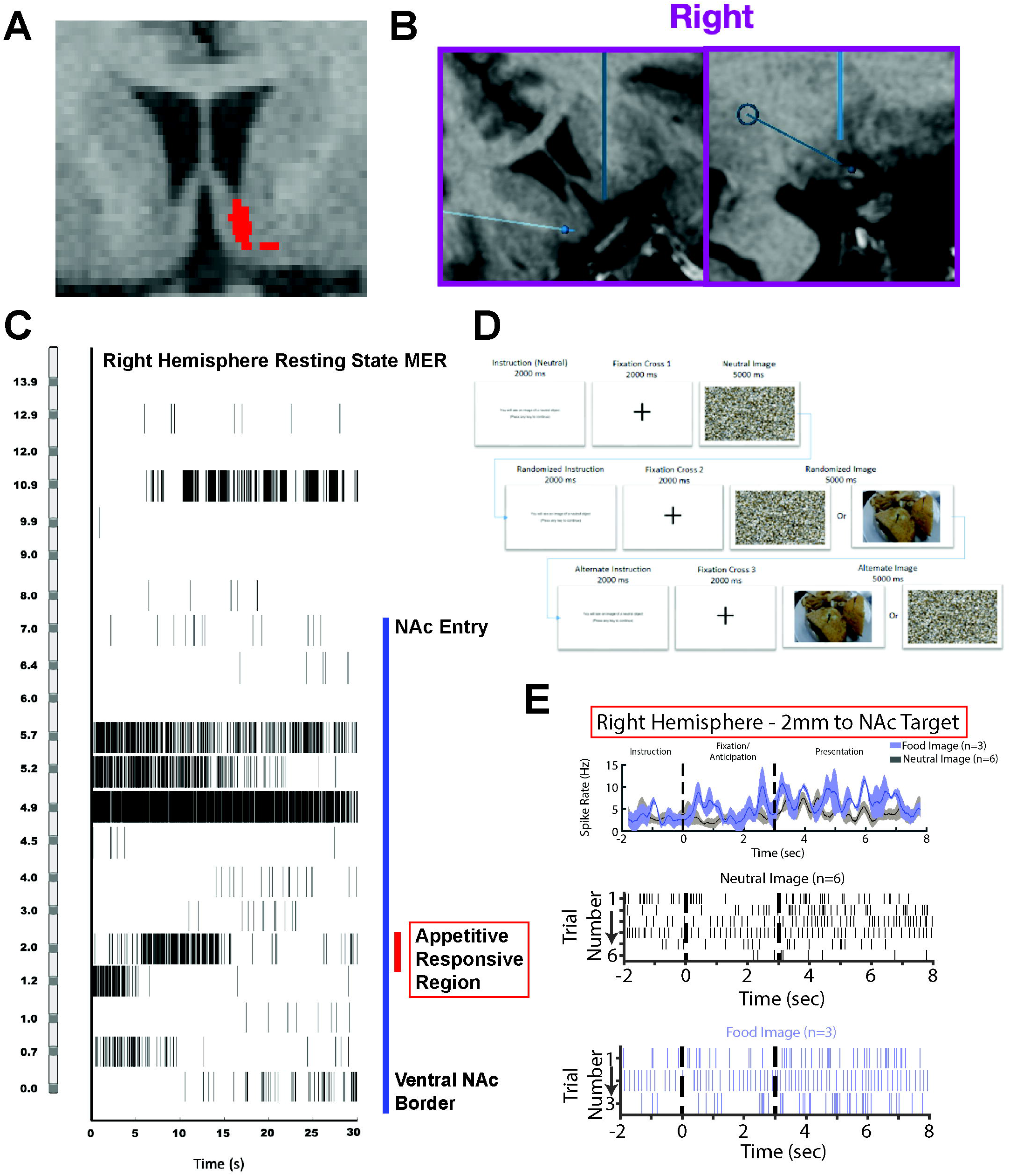
Intraoperative patient-specific appetitive neural activity mapping protocol. A, Red voxels overlaid on the anatomic MR imaging represent the binary cluster NAc subdivision where most of the tractography-defined NAc to LH projections were predicted in the right hemisphere. B, T1-weighted anatomic MR images demonstrate the right hemisphere MER pass trajectory (thick blue line) as captured via intraoperative imaging after the MER pass. Coronal (left) and oblique sagittal (right) reformats through the plane of the microelectrode are depicted, highlighting internal capsule traversal prior to entry into the gray matter of the NAc region. C, Baseline resting-state single unit activity in approximately 1 mm intervals advancing towards the ventral base of the NAc target. Each row represents a distance in millimeters (see y-axis) to the ventral base of the target where the recording was obtained, with raster plot using vertical lines to depict neuronal spike activity. Each baseline recording persisted for 30 seconds (x-axis). The NAc was entered by the microelectrode at approximately 7 mm from the base of the target (blue vertical line), located several millimeters dorsal to the mapped region responsive to appetitive stimuli (red vertical line and box). D, Personalized appetitive visual stimuli were acquired and scored preoperatively. These images were digitally displayed during the performance of an intraoperative clinical instrument designed to provoke anticipation using visual stimuli with time-locked instruction and synchronized microelectrode recordings. E, (Top) Average spike-rate during the provocation using appetitive food and pixel scramble images. Spike-rates are displayed from 2 mm dorsal to the NAc base on the right hemisphere MER pass, the location where the surgical and interventional psychiatry team discerned audibly provocable units in real-time during the provocation instrument administration. (Middle) Raw raster plots of spiking data from the pixel scramble images trials (black) and appetitive food images (bottom) at the same depth along the MER pass. MER, microelectrode recording. NAc, nucleus accumbens.

## Baseline and appetitive neurophysiology of the human NAc region

Reports of high-spatial resolution single-unit and local-field potential recordings of the human NAc region remain sparse (5,8,20–22). Thus, baseline resting state MER along the trajectory (Figure 1B) was recorded with the patient awake in approximately 1 mm intervals (Figure 1C). We initiated appetitive provocation on the right (Figure 1C-D), starting at 7 mm above NAc base. Provocation was initiated, starting at the dorsal border of the NAc visualized on the planned trajectory (Figure 1B-C). The instrument was used approximately every 1 mm and at the discretion of the driving neurosurgeon (CH) if an audibly distinct single or multi-unit activity was detected. The raw MER voltage signal was visually thresholded to generate an auditory spike output for intraoperative real-time interpretation. Starting at this level, audibly discernable differences in activity were present though no correlation to image presentation was appreciated. The MER was advanced until 2 mm above target where there were audible units - the approximate region of the ventral NAc predicted by tractography to contain the highest concentration of LH streamlines. At this depth, provocation was repeated including single presentations of multiple appetitive food items (n=3) and neutral pixel-scramble (n=6) images (Figure 1D). Raw, high-impedance MER signal time-locked with image presentation was processed offline to perform spike sorting and define individual units from which the spiking rate was obtained (Figure 1E). Intraoperatively, this anatomic region exhibited audibly responsive units during appetitive provocation that were otherwise not discerned throughout this pass (Video 1). Intraoperative imaging was performed to confirm the microelectrode trajectory and traversal through the NAc subdivision where most tractography-defined LH projections were localized (Figure 1A-B).

## Intraoperative monopolar stimulation mapping to confirm target engagement

After awake mapping, a quadripolar depth lead (DL-330-3.5K, NeuroPace, Mountain View, CA) was advanced to target. Post-operative imaging demonstrated engagement of the tractography-defined subregion (Figure 2). We anticipated that high-frequency stimulation applied to the NAc region would generate a mirth response or “smile” (23–27). As there have been isolated reports of panic, fear, and anxiety responses during NAc stimulation, we wanted to screen for negative stimulation effects, as their presence if persistent could limit programming. Additionally, prior studies have suggested intraoperative mood response to predict response to DBS (23). We utilized escalating current (0.5 mA to 5.5 mA maximum) with short-burst (10 seconds) monopolar high-frequency stimulation (Figure 2E) performed via a neurostimulator (NeuroPace, Mountain View, CA). After obtaining baseline intraoperative scores for mood, anxiety, and energy levels, stimulation mapping was initiated. Stimulation of the two ventral-most contacts in each hemisphere, those located in the NAc region (Figure 2C-D), demonstrated dose-dependent escalation of mood elevation on a 10-point visual analog mood scale (VAMS) score in accordance with our hypothesis (Figure 2F). The patient’s anxiety score was minimally elevated though with an inconsistent dose response in the ventral most contacts bilaterally (Figure 2F). Self-reported energy was elevated above baseline with current dose dependence and was observed similarly across monopolar stimulation of contacts 1 and 2 bilaterally (Figure 2F). Escalating to currents up to 5.5 milliamperes bilaterally did not reveal any displeasing or depressing phenomena.

**Figure 2.**
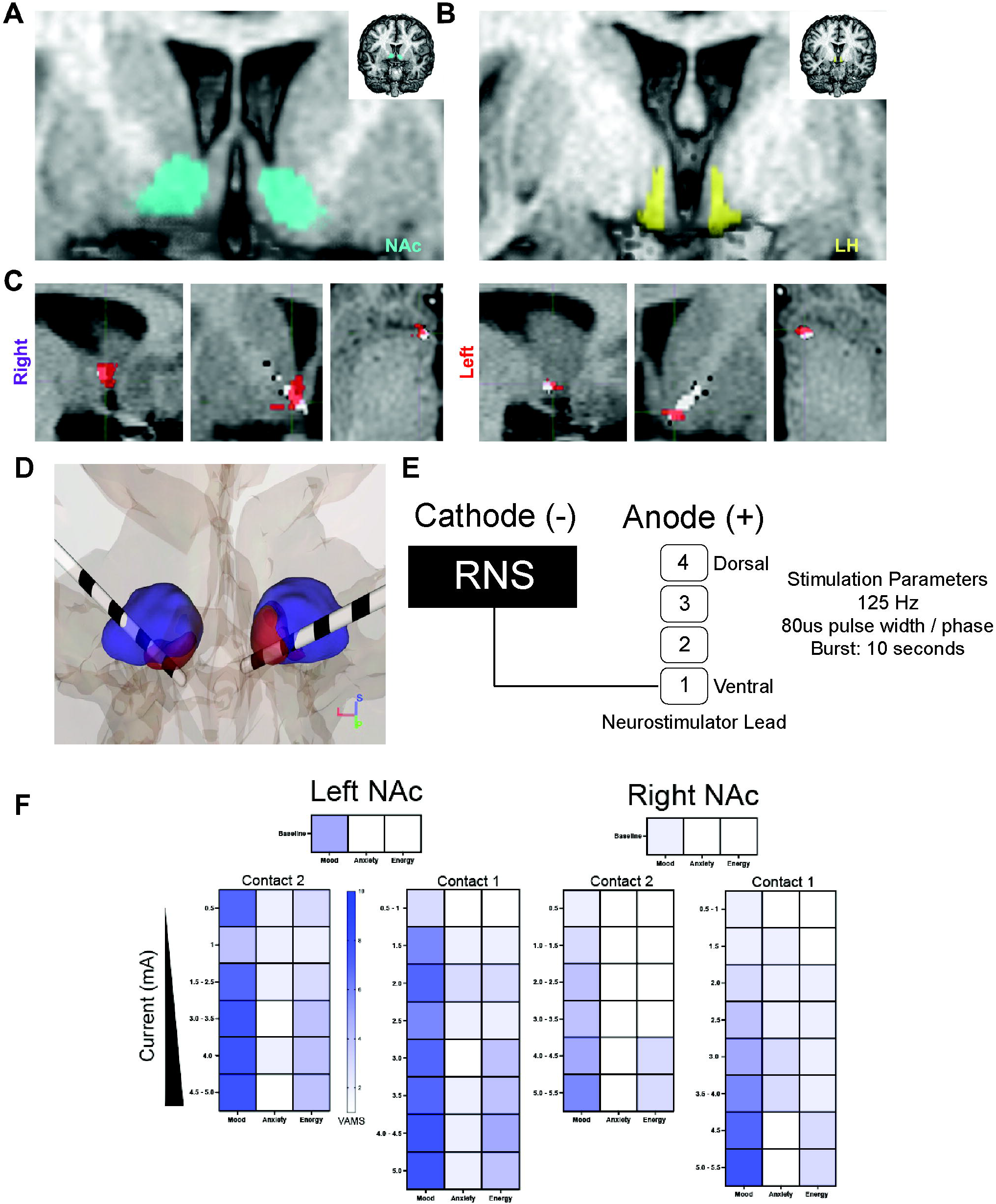
Patient-specific electrode targeting and intraoperative stimulation mapping. A & B, Regions of interest used for probabilistic tractography co-registered to patient-specific anatomical images: NAc (cyan) and LH (yellow), adapted from the CIT168 Subcortical In Vivo Probabilistic Atlas. C, Sagittal, coronal, and axial (left to right) plane T1-weighted MRI reconstructions through the level of the distal tip of the implanted rDBS electrodes (right and left). Electrode position (white pixels) on the postoperative CT was overlaid on the anatomic T1-weighted MR imaging via CT-to-MR fusion (Advanced Normalization Tools and FSLeyes software). Red voxels overlaid on the anatomic MR imaging represent the binary cluster NAc subdivision where most of the tractography-defined LH connections were predicted. The locations of the rDBS electrodes are highlighted within the NAc region informed by tractography-defined interconnections to the LH. D, Three dimensional reconstruction of the final rDBS lead position with respect to the NAc region and predicted LH-interconnected subdivision. The hemispheric asymmetry of the predicted LH projection augmented within the NAc is highlighted and was captured between contacts 1 and 2 of the implanted electrodes bilaterally. E, Standard high frequency 10-second bursts of biphasic charged balanced stimulation pulses were delivered in monopolar fashion using a neurostimulator (NeuroPace, Inc, Mountain View, CA). F, Monopolar stimulation with the parameters depicted in E was performed bilaterally. Pre-stimulation baseline scores in mood, anxiety, and energy were collected prior to stimulation of each NAc region on a 10-point VAMS. Under intraoperative supervision of an interventional psychiatrist (NRW), escalating current dose was utilized during assessment of clinical response in the mood, anxiety, and energy domains. The respective ratings are represented in color temperature along the VAMS. NAc, nucleus accumbens. LH, lateral hypothalamus. rDBS, responsive deep brain stimulation. VAMS, visual analog mood scales.

The ability of tractography-based imaging to reliably predict DBS-mediated clinical outcomes remains under active investigation (13,28,29). Further, the appetitive mapping protocol using a clinical instrument extends our characterization of the electrophysiologic spatial topography and anatomical subregions of the human NAc region (8,11,30). We postulated that the ability to functionally map personalized appetitive responses within the NAc region would provide physiologic confirmation of engagement of a patient-specific tractography-defined circuit. Ongoing use of this instrument will enable prospective study of the topography of response to personalized appetitive stimuli in the human NAc.

## Data Availability

All data produced in the present study are available upon reasonable request to the authors

## Acknowledgements

This work was supported by the National Institute of Health (5UH3NS103446-02). The authors thank the study subject for their dedication and commitment to this novel, first-in-human exploratory trial; Dr. Ian Kratter, Tricia Cunningham, Aroonwan Thongkwan, Vyvian Ngo, and Bharati Sanjanwala for support during surgery and intraoperative testing; Emily Mirro for providing technical support for the NeuroPace neurostimulator. Thanks to Keith Sudheimer for providing the script to assist in generating the scrambled images for the provocation images and Kai Miller for support in neural spiking analyses and video synchronization.

## Author Contributions

J.J.P, D.A.B, C.H, and R.S.S designed experiments, conducted experiments, and conducted analyses. N.W. performed intraoperative psychiatric assessments. J.J.P, C.E.R, D.A.B, C.H, and R.S.S wrote the paper.

---

**Video 1: Intraoperative Audio Recording Demonstrating Appetitive Units within the Human NAc**

Video and accompanying audio demonstrates a representative single-unit appetitive provocation response. Microelectrode recordings were performed 2 mm above the ventral base of the NAc target on the right hemisphere pass prior to RNS quadripolar lead implantation. In this example, the red line moving from left to right marks the time advance over the first 18 seconds of a provocation image trial. The high-impedance microelectrode tip raw voltage, high-pass filtered, and spiking thresholded raster map are shown (top to bottom). The first stimuli depicted here was an image scramble control, followed by presentation of a highly appetitive food stimulus provided by the patient. The anatomic location of these recordings overlapped with the region of increased predicted connectivity from NAc to LH and were noted by the intraoperative team to be an audibly responsive region.

