## Supplementary material for "Appetitive mapping of the human nucleus accumbens": Video 1

### Slide 1
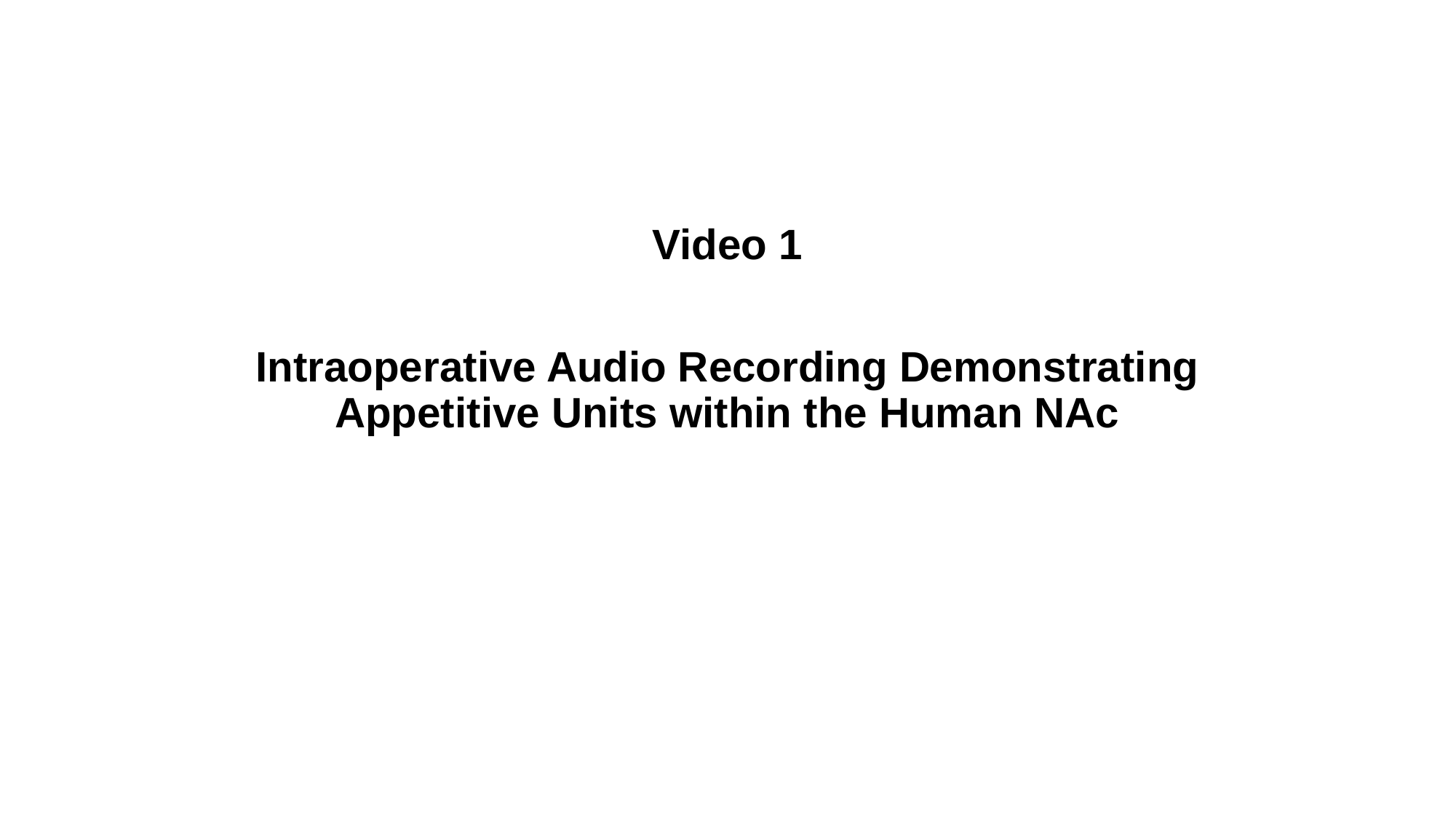

Video 1
Intraoperative Audio Recording Demonstrating Appetitive Units within the Human NAc

### Slide 2
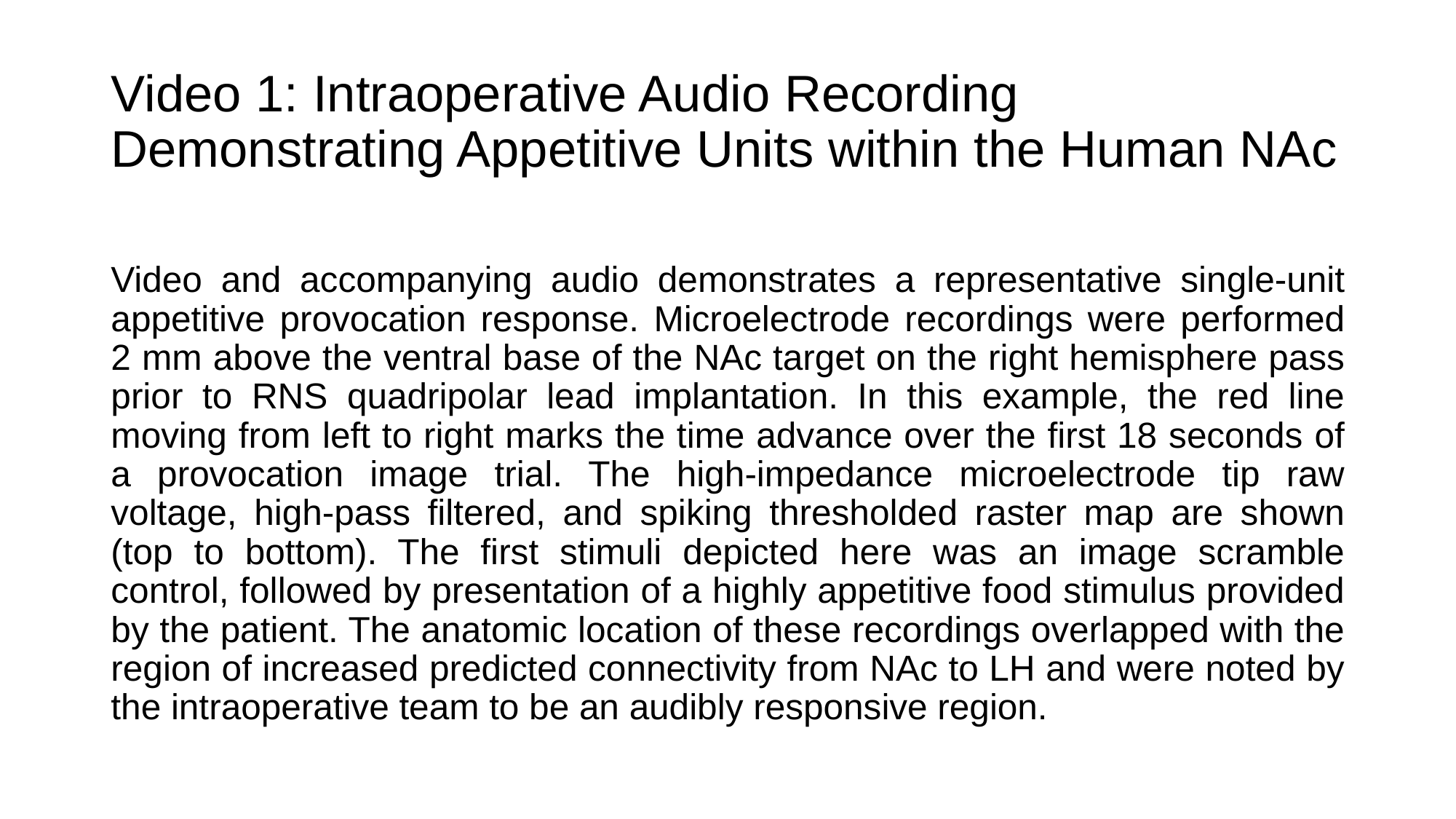

#### Video 1: Intraoperative Audio Recording Demonstrating Appetitive Units within the Human NAc
Video and accompanying audio demonstrates a representative single-unit appetitive provocation response. Microelectrode recordings were performed 2 mm above the ventral base of the NAc target on the right hemisphere pass prior to RNS quadripolar lead implantation. In this example, the red line moving from left to right marks the time advance over the first 18 seconds of a provocation image trial. The high-impedance microelectrode tip raw voltage, high-pass filtered, and spiking thresholded raster map are shown (top to bottom). The first stimuli depicted here was an image scramble control, followed by presentation of a highly appetitive food stimulus provided by the patient. The anatomic location of these recordings overlapped with the region of increased predicted connectivity from NAc to LH and were noted by the intraoperative team to be an audibly responsive region.

### Slide 3
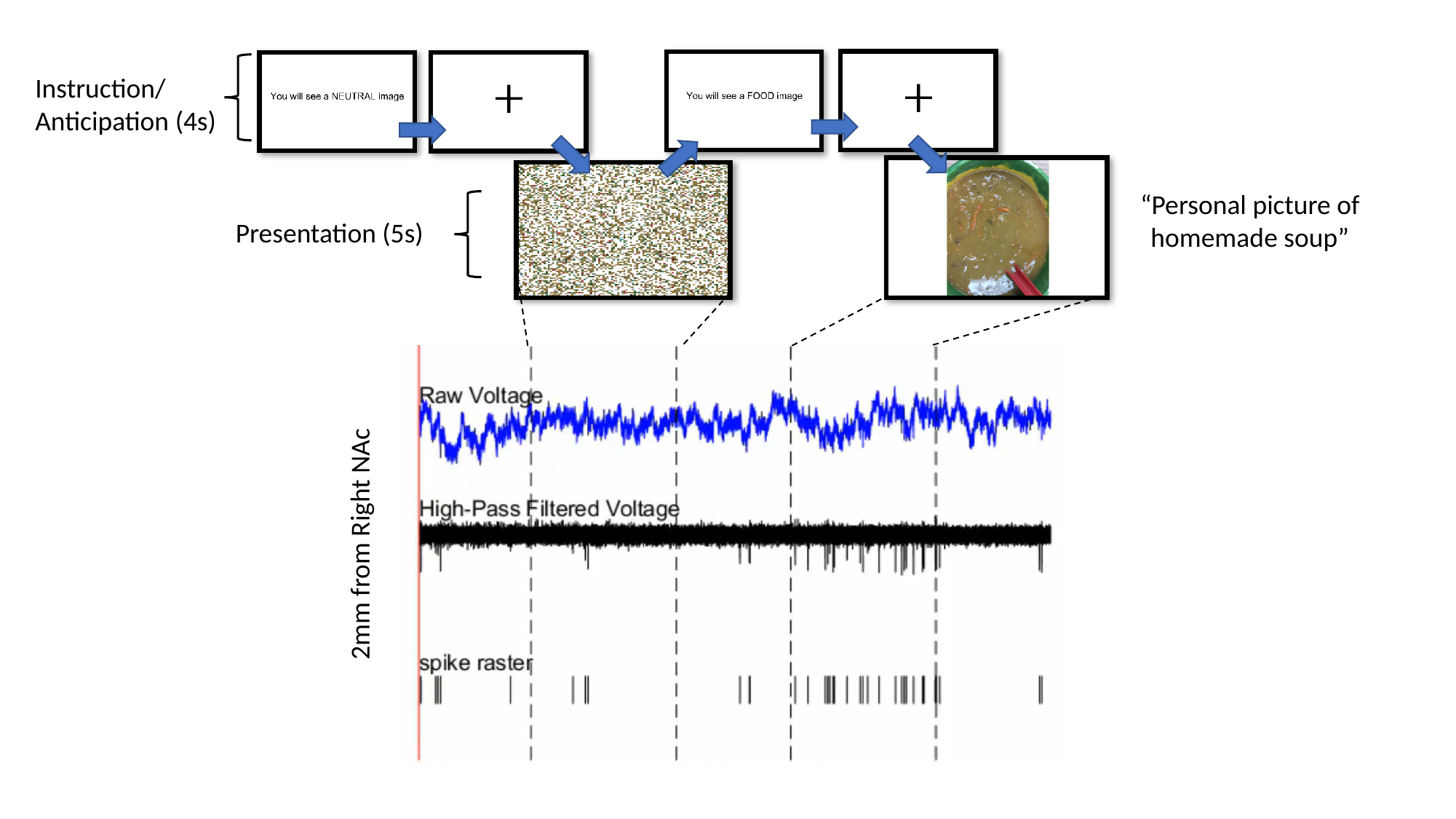

Instruction/
Anticipation (4s)
“Personal picture of homemade soup”
Presentation (5s)
2mm from Right NAc
